# Understanding the role of mask-wearing during COVID-19 on the island of Ireland

**DOI:** 10.1101/2022.03.25.22272946

**Authors:** Nicola Fitz-Simon, John Ferguson, Alberto Alvarez-Iglesias, Mircea T. Sofonea, Tsukushi Kamiya

**Affiliations:** HRB Clinical Research Facility, National University of Ireland Galway, Ireland; MIVEGEC, Univ. Montpellier, CNRS, IRD, Montpellier, France; Center for Interdisciplinary Research in Biology (CIRB), Collège de France, CNRS, INSERM, Université PSL, Paris, France

## Abstract

**Background:** Non-pharmaceutical interventions (NPI) play a key role in managing epidemics, yet it is challenging to evaluate their impacts on disease spread and outcomes.

**Methods:** To estimate the effect of a mask-wearing intervention to mitigate the spread of SARS-CoV-2 on the island of Ireland, we focused on the potential for interindividual infectious contact over time as the outcome. This is difficult to measure directly; in a companion paper we estimated it using a multi-strain epidemiological model. We used data on mask-wearing and mobility in both Northern Ireland (NI) and the Republic of Ireland (ROI) to predict independently the estimated infectious contact over time. We made counterfactual predictions of infectious contact rates and hospitalisations under a hypothetical intervention where 90% of the population were wearing masks during early 2020, when in reality few people were wearing masks in public; this was mandated in both jurisdictions on 10th August 2020.

**Results:** There were 1601 hospitalisations with COVID-19 in NI between 12th March and 10th August 2020, and 1521 in ROI between 3rd April and 10th August 2020. Under the counterfactual mask-wearing scenario, we estimated 512 (95% CI 400, 730) hospitalisations in NI, and 344 (95% CI 266, 526) in ROI, during the same periods.

**Conclusions:** We have estimated a large effect of population mask-wearing on COVID-19 hospitalisations. This could be partly due to other factors that were also changing over time.

## Introduction

Non-pharmaceutical interventions (NPI) are actions taken by individuals or communities that aim to reduce infectious contacts between susceptible and infectious people during an epidemic [1]. Often governments may mandate such actions. Examples include reducing the number of physical contacts (e.g., closure of schools, workplaces, commercial establishments, roads, and public transit; restriction of movement; cancellation of public events; maintenance of physical distances in public), reducing the chance of infection upon contact (e.g., mandatory use of personal protective equipment), and identifying and isolating those that may be infected (e.g., contract tracing and digital surveillance). By slowing the surge of infection, communities are afforded an opportunity to reduce infection-induced mortality and morbidity, alleviate health care burden and wait out an epidemic until pharmaceutical solutions (i.e. treatment and vaccines) become available. Public health policies to reduce the mixing of susceptible and infectious people have been instrumental in historical outbreaks, including during the 1918 influenza pandemic where rapid implementation of NPI mandates was crucial for reducing excess death in the United States [2].

Due to the rapid spread of SARS-CoV-2 and the initial lack of effective therapy, NPI have been central to managing the COVID-19 pandemic globally. NPI such as staying at home and mask-wearing, both of which are often mandated, can reduce the reproductive number (i.e., the average number of secondary cases per infectious host) and sometimes bring it below 1, thereby halting the growth of SARS-CoV-2 infections in a population [3, 4]. Reducing inter-individual infectious contact has also been shown to reduce the relative advantage of highly transmissible variants [5]. Even after a widespread rollout of vaccines, understanding the effectiveness of NPI and the role of mandating them remains pertinent as vaccination alone is unlikely to put an end to the pandemic [6] and vaccination and NPI can synergistically reduce SARS-CoV-2 transmission [7]. Despite their public health benefits, social distancing measures have been shown to incur high costs in several domains, including in the economy [8], mental health [9], and civil liberty [10]. Thus, it is crucial to quantify the effectiveness of interventions to achieve desired public health outcomes and improve policy transparency and public engagement.

Mechanistic epidemiological models have been widely applied to study the dynamics of SARS-CoV-2, and to make predictions of clinical outcomes under alternative scenarios (e.g. an assumed decrease in physical contact [11]). Furthermore, fitting a mathematical model to data on observed processes such as reported cases and hospital admissions allows estimation of inter-individual infectious contact [12, 13]. In a companion paper, we have estimated longitudinal infectious contact ratios for SARS-CoV-2 in Northern Ireland (NI) and the Republic of Ireland (ROI) using a multi-strain compartmental model ([14]). While mechanistic epidemiological models are not primarily intended for the estimation of the effects of NPI, there has been some recent work on incorporating effect estimation in a mechanistic or semi-mechanistic modelling framework. For example, the Institute for Health Metrics and Evaluation (IHME) COVID-19 Modelling Team used compartmental epidemiological models to estimate time-varying rates of infection of susceptibles, and subsequently used regression models to predict these rates in terms of NPI variables; this allowed them to make forecasts of infection rates, hence mortality and morbidity outcomes [15]. Other relevant approaches that link NPI to disease transmission include those that incorporate digital mobility data in a (semi-)mechanistic disease model (e.g., [16, 17]). An alternative approach involves structural equations that model potential outcomes such as deaths or cases in terms of interventions of interest (e.g., [18–20]).

Our aim in this paper was to investigate outcomes under a counterfactual scenario wherein most people wore masks in public during the early part of the COVID-19 epidemic in Ireland, which included the first wave. In reality, public health advice discouraged population mask-wearing during the early part of the epidemic, and few people wore masks in public places during the first wave [21]. We compiled and described data on actions that aimed to reduce the potential for inter-individual infectious contact on the island of Ireland, including relevant data on behavioural changes such as mobility patterns and mask wearing prevalence. We used these data to independently predict infectious contact ratios we had previously estimated for both jurisdictions on the island of Ireland ([14]). We made counterfactual predictions of infectious contact and ensuing hospitalisations under a hypothetical intervention where 90% of the population were wearing masks in public during the first six months of the epidemic in Ireland.

## Methods

### The infectious contact ratio

The infectious contact rate at any point in time is a measure of the potential for inter-individual infectious contact; we estimated this as a ratio relative to its value at the beginning of community spread in Ireland, which we previously estimated to be around 5th March 2020 ([14]). To estimate longitudinal infectious contact ratios, we used a discrete-time compartmental model that incorporates multiple virus strains with different transmissibilities. We estimated these by week for both NI and the ROI over the first year of the pandemic (5th March 2020-28th February 2021), fitting the model to longitudinal data on hospital admissions with COVID-19 and proportion of B.1.1.7, or Alpha, strain cases in each jurisdiction. To parameterise the model, distributions of the time from infection to transmission, and from infection to hospitalisation for hospitalised cases, were based on published evidence [22–24]. As our epidemiological model allowed for durations in each disease compartment informed by evidence, we were able to model accurately the short-term dynamics of the disease, meaning that the infectious contact ratios we estimated reflect short-term behaviour. This makes the infectious contact ratio a suitable outcome for modelling in terms of predictors that also changed in short time frames, such as mobility. The model, model fitting in a Bayesian framework, and results are described in detail in our companion paper [14]. For subsequent analyses in the current paper, we took a single replicable draw from the posterior distribution of estimated weekly infectious contact ratios over the first year of the pandemic for each jurisdiction, and used the posterior medians as our measure of infectious contact.

### Data to predict infectious contact ratio

The governments of ROI and NI carried out a series of public health interventions to curtail the transmission of SARS-CoV-2. We used publicly available sources to compile data on timing and type of policy restrictions in NI the ROI between March 2020 and March 2021, further detail on these is available in [14]. NI followed a different approach to the ROI in introducing and easing restrictions; the UK has a COVID-19 alert system with five levels of alert depending on the level of COVID-19 in the community [25], whereas the ROI defined levels of restriction to mitigate the impact of the virus [26].

As a measure of the extent of physical contact between people, we extracted data from the COVID-19 Google Community Mobility reports for NI and the ROI. These provide six population mobility streams relative to a pre-pandemic baseline [27]. We computed a seven-day moving average of three streams, namely workplaces, transit stations and retail and recreation, as these are thought to be the most appropriate to reflect relevant mobility patterns ([28] used an average of these three streams plus grocery and pharmacy to reflect relevant movement patterns outside the home). As the mobility data for NI were reported by local government district, we calculated an overall seven-day moving average weighted by the population of each district. Further detail on the mobility streams is provided in Supporting Information S1.

We gathered data on the population proportion reporting to be wearing masks in public, and other behavioural data, from behavioural surveys published by the Irish Department of Health, and the Northern Ireland Statistics Research Agency (NISRA) [29,30]. These were based on regular cross-sectional behavioural surveys of 1200 people in NI and 1500-2000 in the ROI; the NI samples were weighted for household size and nonresponse and the ROI samples weighted to demographics [29, 31]. While these behavioural surveys were done from March 2020 in ROI and April 2020 in NI, questions about mask use in public places were only included from 2020-05-04 in the ROI, and from 2020-06-17 in NI. We assumed 1% of the population were wearing masks in public at the beginning of the epidemic on the island of Ireland, and fit a binomial regression model with a spline smooth for time to predict proportion wearing masks on dates not reported in the surveys.

Seasonality may play a role in virus transmission, as people are more likely to be in indoor settings more conducive to disease spread during colder months [32]. To define seasons, we used Met Éireann daily weather data from Dublin airport [33]. We calculated 7-day running means of the median daily temperature, and defined the summer season to start when this temperature first exceeded 10.5 degrees centigrade, and to end when it first dropped below this value. The summer season thus defined ran from 17th May until 21st September. Using warmer temperatures to define the summer period aims to identify a period when people are less likely to be indoors.

### Regression models

We described the relationship between the estimated weekly infectious contact ratios, the overall Google mobility stream (modelled linearly), and longitudinal mask-wearing proportion in each jurisdiction. We fitted a regression model with response the natural log of the weekly contact ratios and predictors the jurisdiction (NI or ROI), the season (summer or winter), the Google mobility stream, and smoothed proportion mask-wearing at the mid-point of each week. We assumed the effect of mask wearing conditional on the other predictors to be linear and the same in each jurisdiction. We included an interaction term between season and proportion mask-wearing to allow for a different effect of mask-wearing in the winter when people are more likely to be indoors. We included an interaction term between mobility and mask wearing to allow for a change in the effects of each as the level of the other changes. For instance, the effect of increasing mobility may be attenuated when most people are wearing masks compared to when few people are wearing masks. To explore assumptions of linearity, we also fit a model including spline smooths for the effect of the proportion wearing masks on the response. We calculated goodness-of-fit statistics for the predictive models.

Our regression model assumes that the responses are independent, which is not the case, as they are correlated over time due to the smoothing described in Kamiya et al. [14]. Therefore, we estimated heteroskedasticity and autocorrelation consistent Newey-West standard errors for the parameter estimates, hence adjusted the confidence intervals for the predicted responses from separate models for each jurisdiction [34].

### Counterfactual scenarios

We compared the observed responses with predicted responses under a hypothetical intervention where 90% of the population were wearing masks during the early months of the pandemic. In effect, we leveraged observed data from late 2020, when over 90% of the population were wearing masks, to estimate the counterfactual scenario.

We checked that the number of hospitalisations predicted by the epidemiological model under no intervention was consistent with the observed number of hospitalisations. We then used our multi-strain epidemiological model to predict hospital admissions from the counterfactual infectious contact ratio. We compared the sum of predicted hospital admissions under the intervention with observed hospital admissions up to the date of the mask mandates (2020-08-10 in both jurisdictions).

We estimated confidence intervals for the difference in hospital admissions for the counterfactual scenario versus reality in each jurisdiction by bootstrapping the process of estimating regression models, predicting the outcome under the counterfactual scenario, and using this predicted outcome (the counterfactual infectious contact ratio) to predict counterfactual hospital admissions from the multi-strain epidemiological model. We ran 10,000 bootstrap replications, and estimated bias-corrected and accelerated (BCa) confidence intervals.

We considered the possible impact of confounding by including potential measured confounding variables that were also changing over time in the regression models, and considering the possible impact of unmeasured confounding.

## Results

### Estimated infectious contact and mobility

Infectious contact is likely affected by the extent of physical contact between people, which may be reflected by digital mobility data. To better understand how mobility behaviour affects infectious contact, we explored the relationship between the infectious contact ratio and the overall Google mobility data stream described above. We observed that the relationship between infectious contact and mobility shifted in early summer 2020 and again in January 2021 (Figures 1 & 2). This finding mirrors that of Nouvellet et al [28] who demonstrated a change in the relationship between mobility and the reproductive number in mid-2020 in many countries, which they attributed to the increased use of NPI, such as contact tracing. We also note that the change in the relationship between infectious contact ratio and mobility in both NI and the ROI coincides with an increase in mask wearing (see Figure 3). A later change in the relationship, in January 2021 may be due to the beginning of the vaccination programme (and this effect appears larger in NI, where the vaccination roll out was faster; albeit few second doses were administered during the period we study [35, 36]).

**Figure 1:**
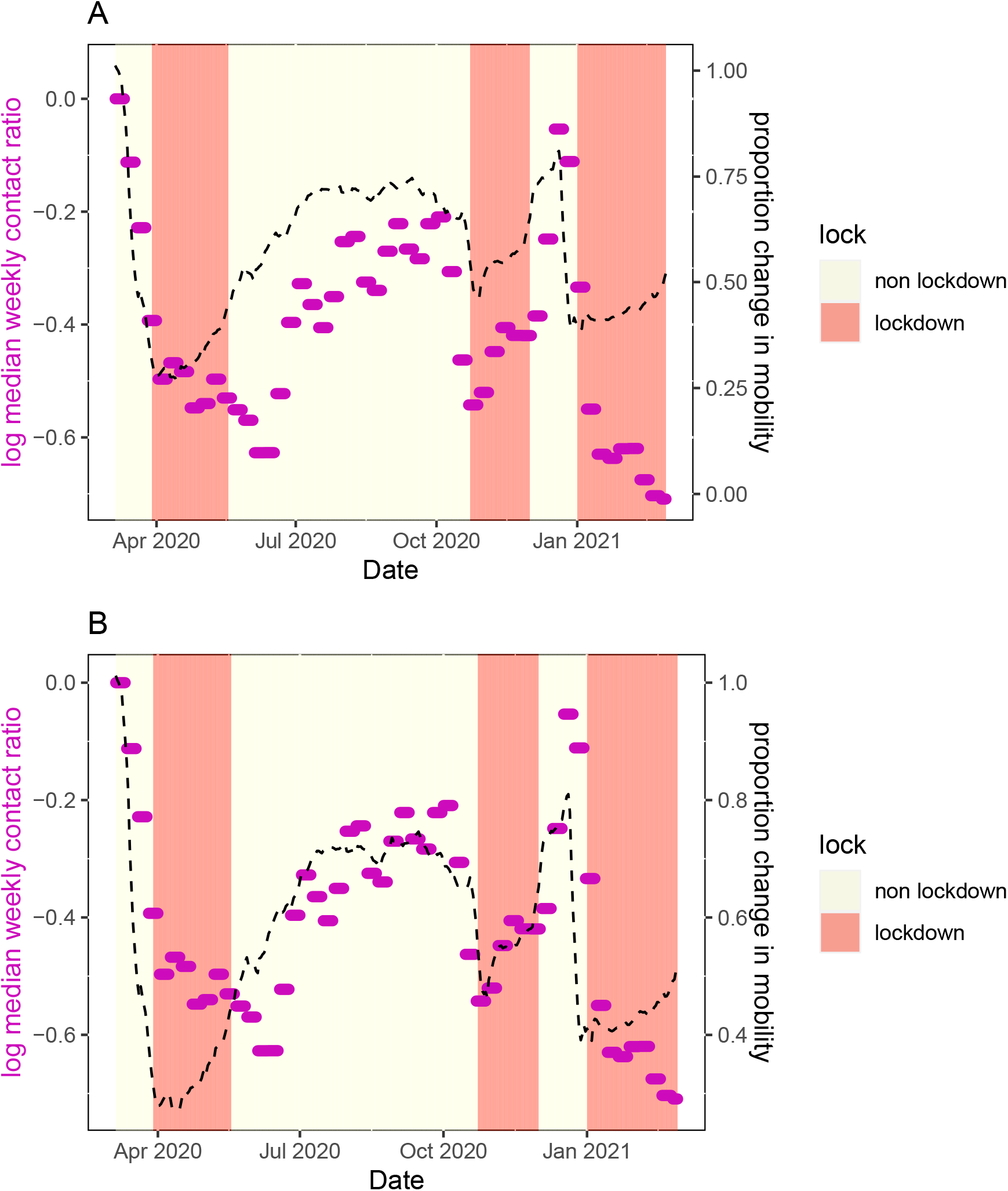
Restriction level, mobility and contact ratios for the Republic of Ireland. Red shaded regions show periods of the highest restrictions (i.e. lockdown). Median weekly contact ratios are shown as purple dots. The averaged Google mobility stream is shown as a black dashed line. In Panel A, the mobility stream is scaled to coincide with contact ratios at the beginning of the period (up to mid May). In Panel B, the mobility stream is 7 scaled to coincide with period May-December.

**Figure 2:**
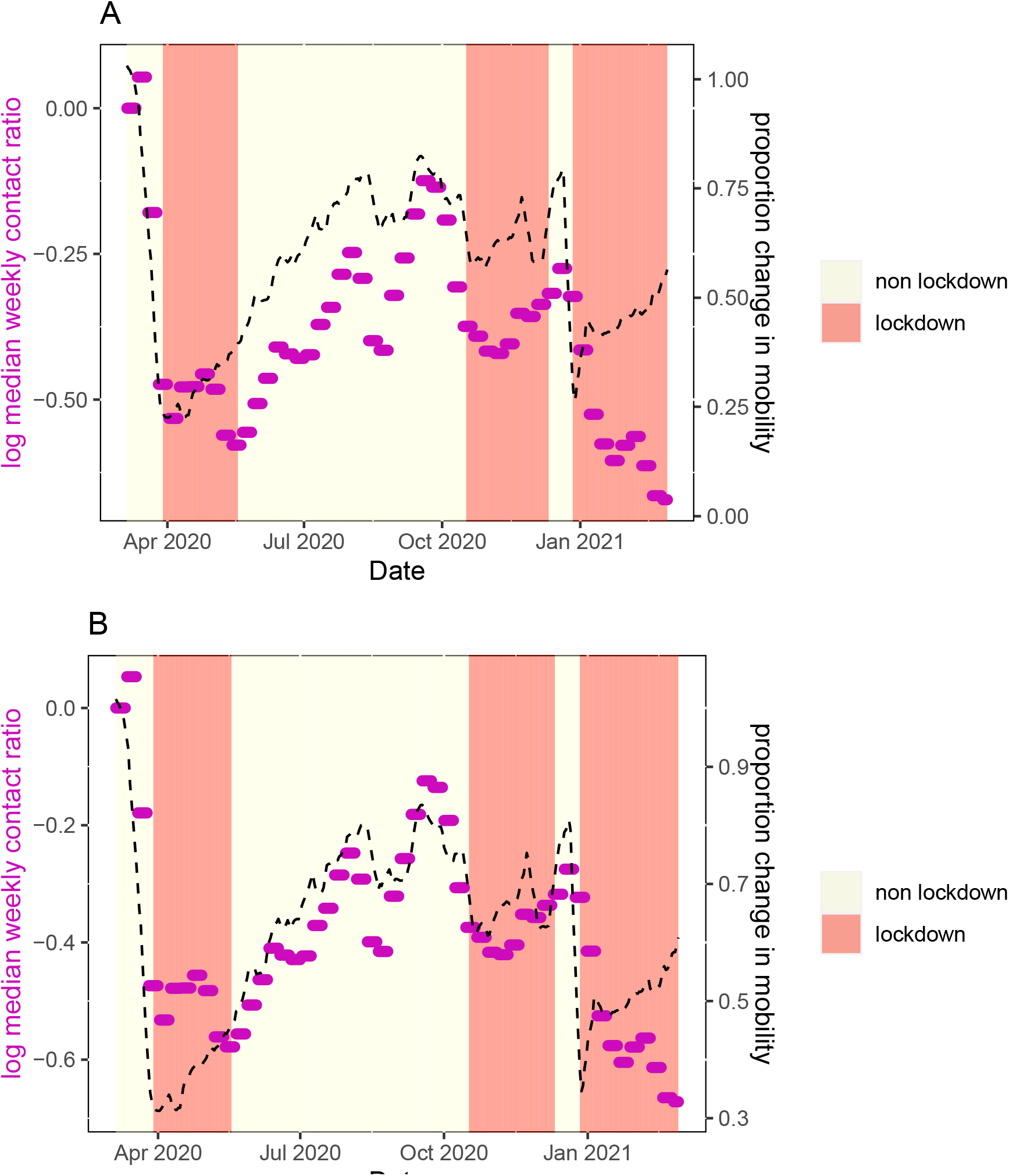
Restriction level, mobility and contact ratios for Northern Ireland. Red shaded regions show periods of the highest restrictions. Median weekly contact ratios are shown as purple dots. The averaged Google mobility stream is shown as a black dashed line. In Panel A, the mobility stream is scaled to coincide with contact ratios at the beginning of the period (up to mid May). In Panel B, the mobility stream is scaled to coincide with period May-December.

**Figure 3:**
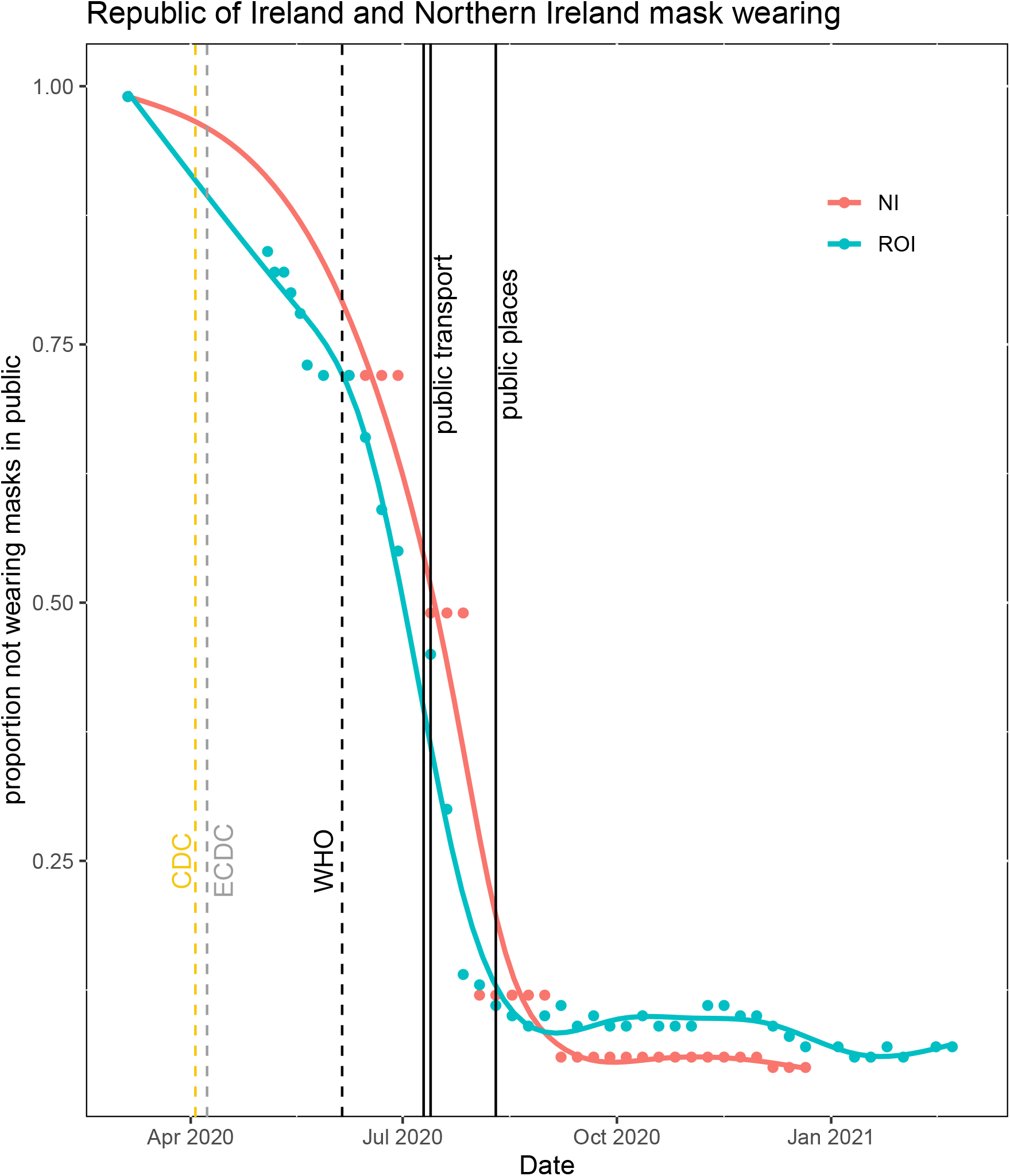
Reported use of face masks in public places on the island of Ireland, with spline smooths for each jurisdiction. Dates of public health mandates to wear masks on public transport (2020-06-10 in NI, 2020-06-13 in ROI), and in all public places (2020-08-10 in both NI and ROI) are shown as black vertical lines. The dates on which major public health institutions changed their recommendations on mask wearing for the general population from advising against wearing masks to wearing masks in indoor public places are shown as dashed vertical lines (US Centers for Disease Control and Prevention, CDC (2020-04-03), European Centre for Disease Prevention and Control, ECDC (2020-04-08), World Health Organisation, WHO (2020-06-05).

### Mask mandates and mask-wearing behaviour

Google mobility data and our epidemiological model demonstrate that decreases in mobility (and infectious contact ratios) preceded lockdowns (Figures 1 & 2). Change in behaviour ahead of government mandates is also evident from comparing mask mandates with level of self-reported mask wearing in public places (Figure 3). In both NI and ROI, around 90% of people reported wearing masks before the date of the national mask mandates on 2020-08-10. While early advice largely discouraged people from wearing masks (e.g., [37]), the US Center for Disease Control and Prevention changed their advice to recommend mask wearing in public places on 2020-04-03 [38], closely followed by the European Centre for Disease Prevention and Control on 2020-04-08 [39]. While behavioural data on mask wearing in public was only collected from May in the ROI and from June in NI, 16% of people in the NI reported wearing masks in early May, and the proportion of people wearing masks increased more rapidly after the World Health Organisation changed their advice to recommend them on 2020-06-05 [40]. Taken together, global and international public health advice likely had a greater influence on the masking-wearing behaviour than the subsequent national mandates in both ROI and NI. Thus, we focus our attention on estimating the impacts of actual behavioural change, in particular mask wearing, rather than the impacts of government mask-wearing mandates.

### Regression models to predict infectious contact ratio

We fit regression models for 2020 data only, as we have observed that the relationship between our response variable and mobility changes around the end of 2020, around the time the vaccination programme began, and it is possible this would have impacted on the potential for inter-individual infectious contact. We found that the regression model with a linear effect of proportion wearing masks predicts the log weekly contact ratios reasonably well (Figure 4; broken lines); with AIC -208.6 and *R*^2^ 0.77. A spline smooth for mask-wearing provides a better fit with AIC -218.1 and *R*^2^ 0.81. However the improvement is not vast, and the assumption of a linear relationship with log contact ratio is more consistent with other evidence about population mask-wearing [41]; we therefore assume a linear effect for the subsequent counterfactual analysis.

**Figure 4:**
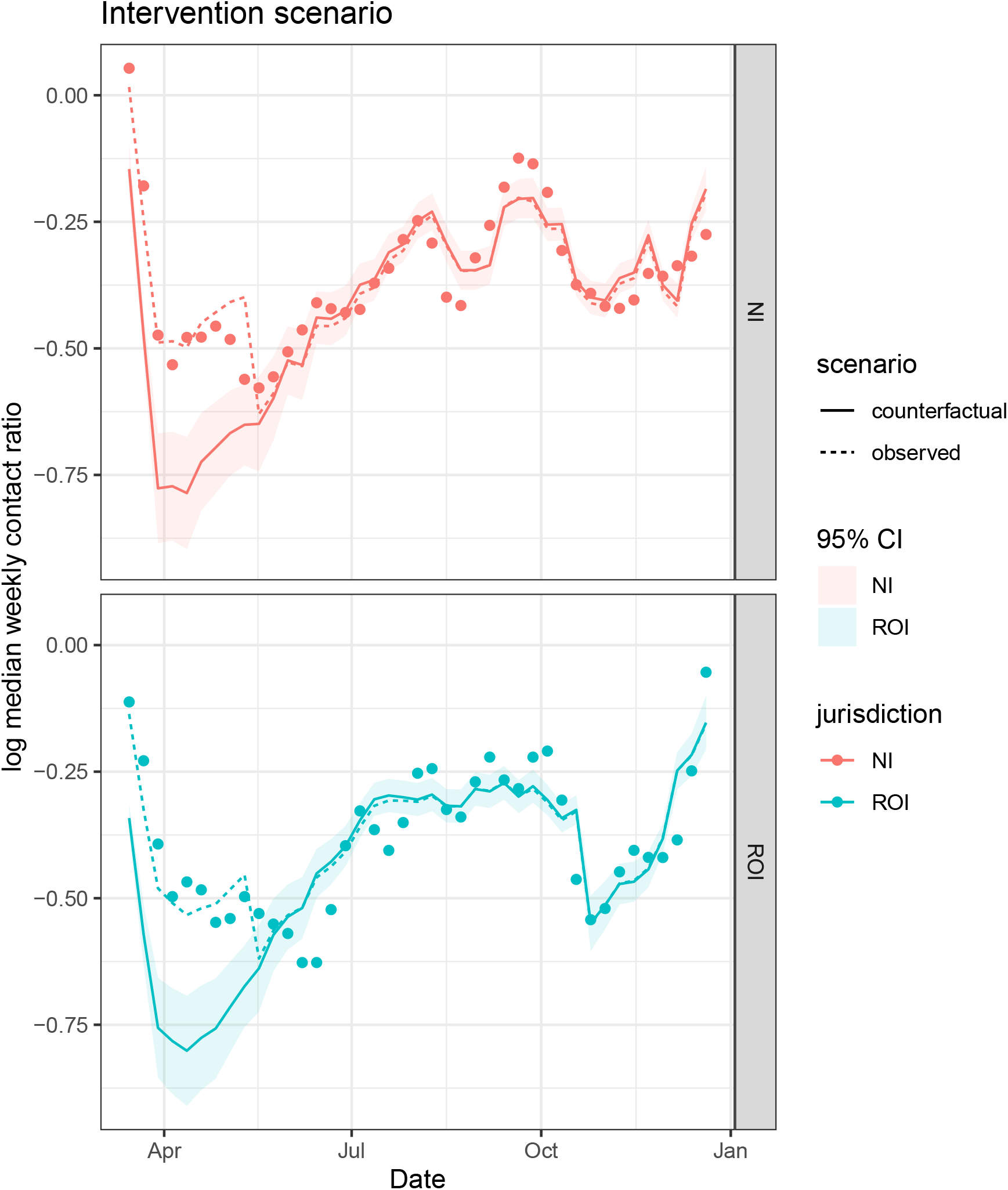
The dots show the log weekly contact ratios. The dotted lines are the predicted log contact ratios from a linear regression model with predictors jurisdiction, season, average mobility and proportion of people wearing masks. The solid lines show predicted log contact ratios and 95% CIs under a hypothetical intervention where 90% of people wore masks throughout. In the hypothetical situation, mobility is assumed to remain as observed in reality.

Newey-West heteroskedasticity and autocorrelation consistent sandwich estimators for the parameter variances in regression models for each jurisdiction separately gave slightly decreased standard errors of most parameter estimates [34]. This deflated pointwise confidence intervals for predicted responses slightly in some regions, and inflated them slightly in others (See Supporting Information S2). As the impact on the confidence bands for predicted log infectious contact was small, we ignored the non-independence of responses when estimating confidence intervals for the number of hospitalisations saved.

### Counterfactual scenarios

We used the linear regression model to predict log infectious contact under the counterfactual scenario that 90% of the population were wearing masks during the early part of the epidemic on the island of Ireland up to the date of the mask mandates, while mobility remained as observed in reality. The counterfactual scenario predicts a marked decrease in infectious contact during the first wave in both jurisdictions compared with that observed in reality (Figure 4; solid lines). Supporting Information S3 shows a contour plot of observed data together with the model predictions as a function of mobility and mask wearing for summer and winter separately. We have considered an intervention on mask wearing for which we have a reasonable amount of observed data to estimate the counterfactual scenario. However this does involve some extrapolation beyond the range of the observed data to a region of high mask wearing and very low mobility.

While there were in total 1,601 observed COVID-19 hospitalisations in NI from 12th March until the date of mask mandate on 10th August (1690.3 predicted from epidemiological model), we predicted 512.0 (95% CI 399.6, 729.8) under the counterfactual scenario - that is 1089 (871,1201) fewer hospitalisations. Daily hospital admissions in the ROI were not publicly available before 3rd April, but there were 1,521 admissions between this date and the mask mandate (1531.8 predicted from the epidemiological model), while our counterfactual mask-wearing scenario predicted 343.8 (95% CI 266.2, 526.2) between 3rd April and 10th August - that is 1177 (995, 1255) fewer hospitalisations.

The behavioural data reported by the Irish Department of Health also included information on other NPI measures: the proportions of people reporting to be sitting further apart from others, and distancing when in a queue, compared to before the pandemic, and the proportions of people washing their hands, and using sanitiser [29]. We estimated the predicted proportions of these NPI variables from binomial regression models with spline smooth for time, as we did to estimate the predicted proportion wearing face masks at intermediate dates. As a sensitivity analysis for confounding by other NPI variables, we fit regression models for the log weekly contact for the ROI as above, and also adjusting for predicted value of each NPI variable, and an interaction between mask-wearing and the other NPI variable to reflect the assumption that the effects of multiple NPI are not additive. Figure 5 shows for each NPI variable that the predicted log contact ratio under the 90% mask-wearing intervention with adjustment for the NPI variable, was only slightly different than without each adjustment. However, pointwise 95% confidence intervals were greatly inflated for each adjustment apart from hand-washing, so that the actual log contact ratios fell within the adjusted confidence bands. The proportion hand washing was high and fairly constant throughout the period considered; the other three NPI variables were increasing at the same time as mask wearing, though none to the same extent. The inflation of pointwise confidence intervals could be due to co-linearity of the predictor variables.

**Figure 5:**
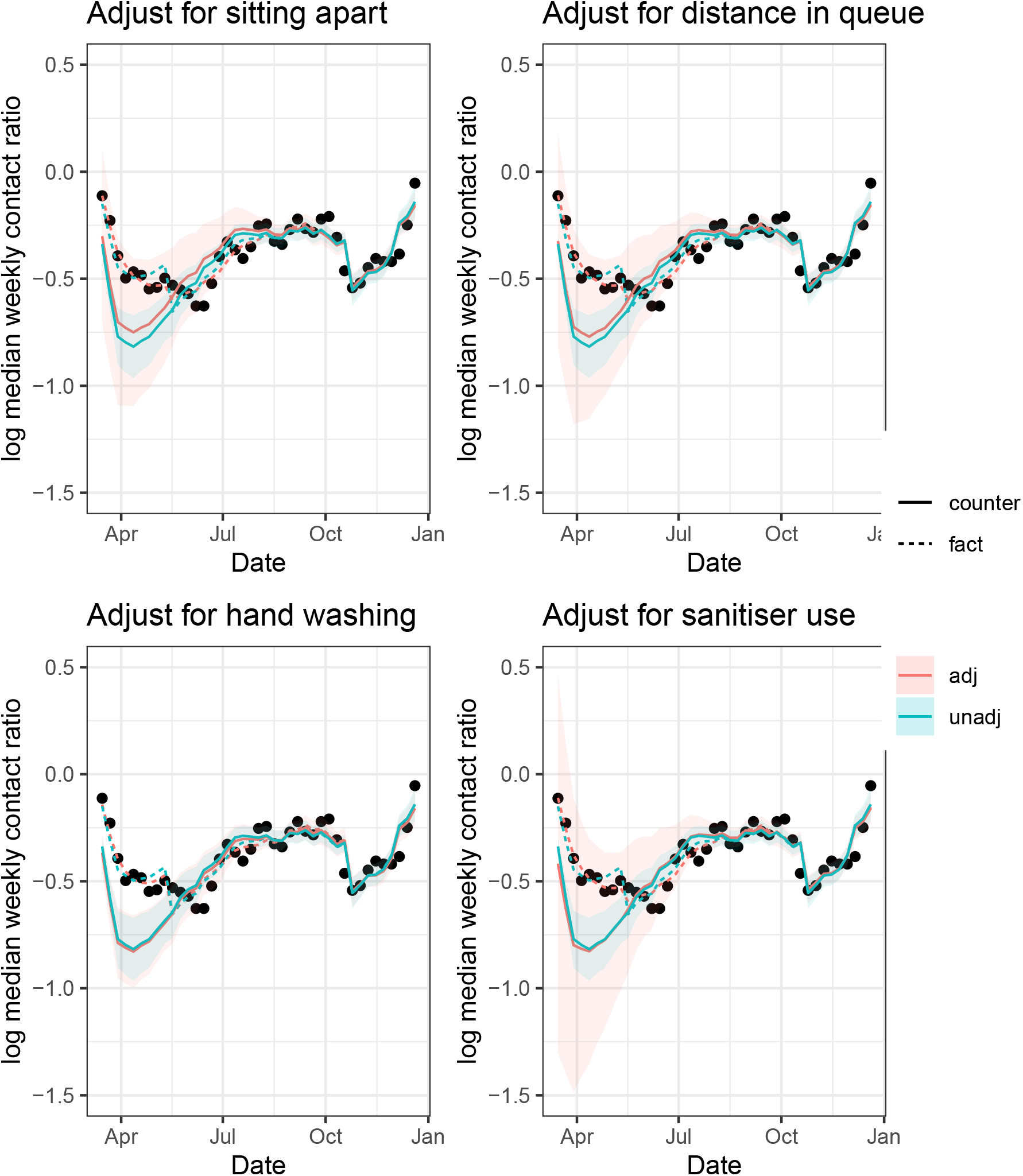
Republic of Ireland: The dots show the log weekly contact ratios. The dotted lines are the predicted log contact ratios from a linear regression model with predictors season, average mobility, proportion of people wearing masks, and the NPI variable of the title. The solid lines show predicted log contact ratios and 95% CIs under a hypothetical intervention where 90% of people wore masks throughout. In the hypothetical situations, mobility and the other NPI variable are assumed to remain as observed in reality.

## Discussion

Most attempts to estimate the effects of non-pharmaceutical interventions on COVID-19 outcomes — either by incorporating effect estimation in a mechanistic epidemiological model, or by using structural equations models — have been in a US setting (e.g., [15–20]).

In the Irish context, compartmental models been used by several other research groups to understand the dynamics of the virus, make forecasts of outcomes under various scenarios, and assess economic impacts of policy restrictions. For example, the Irish Epidemiological Modelling Advisory Group fit compartmental models to case data, and forecast scenarios under different values of the reproductive number [42]. Ó Náraigh and Byrne [43] applied optimal control theory to data on cases and deaths, and found the optimal strategy to control the epidemic required large and early mitigation strategies. Cazelles et al. [44] used a stochastic model with time-varying parameters and studied the prevalence and tracked the effective reproductive number and its percentage reduction between restriction periods during the first year of the epidemic. Jaouimaa et al. [45] incorporated an age-structured contact matrix to their compartmental model and explored the evaluation of an age-related economic cost of different lockdown measures in the ROI. Several UK studies also included NI data to understand regional COVID-19 dynamics [46, 47].

An important difference between the infectious contact parameter and time varying reproductive number is that the former is unaffected by changes in virus transmissibility, which is modelled independently in our approach [14]. This separation is especially useful in estimating the effects of NPI (such as mask wearing) that might be expected to have similar effects on infectious contact regardless of the variant (or mixture of variants) currently dominant in the population. In contrast, biases may result from estimating the effect of NPI on the time varying reproductive number during transition periods where this mixture in the population is changing. Taking advantage of the estimated infectious contact ratio, we examined its relationship with mobility and mask-wearing.

A further advantage of using the infectious contact ratio as the outcome in our investigation of the effect of population mask-wearing is that the proportion wearing masks may affect the infectious contact ratio at the current week, but should not affect the infectious contact ratio at other (future) weeks. In infectious disease studies, estimating the effect of interventions is challenging when the intervention on one unit can affect the outcome of another, as could be the case if the outcome were the level of infection in the community. Our use of the infectious contact ratio bypasses this problem of interference, with the caveat that there is autocorrelation between successive estimated contact ratios due to the smoothing described in Kamiya et al [14]. Nonetheless, we also found in that study that the bias was small and unlikely to have a substantive effect on our results.

In estimating the effect of mask-wearing on the infectious contact ratio, we have not adjusted for all potential sources of confounding. For example, other actions to mitigate the spread of the virus may also have been changing at the same time as mask wearing, and may be partly responsible for the effect we attribute to wearing masks. Behavioural survey data from the Irish Department of Health and NISRA indicate that many of the relevant behaviours that could bias the effect remained fairly constant throughout (for example hand washing, which was high from early during the period considered), or increased to a smaller extent than mask-wearing (eg self-reported social distancing) [29] [30]. When we included other NPI variables in the regression model, counterfactual predictions were little changed, but confidence intervals were inflated; this could be due to co-linearity.

It is possible that early adopters of mask wearing were those who were at higher risk of severe COVID-19, so that the observed effect of mask wearing reflects the patterns of uptake in the population, and cannot be interpreted as the effect of adoption by a random proportion of the population. We do not have demographic information on the mask-wearers, so cannot adjust for demographic variables that predict COVID-19 severity, such as average age of mask wearers at each time. Over time, people could have been becoming better at taking various different actions to mitigate the spread of the virus, and we have no measure of improvement in implementation of NPI.

As we did not observe many combinations of mask-wearing and mobility, especially when mobility was very low, the counterfactual scenario involves some model extrapolation. This influenced our modelling assumption that the effects of mask wearing on log contact ratio are linear rather than using the better fitting spline smooth; the latter is consistent with the evidence that the the population proportion not wearing masks has a multiplicative effect on the reproductive number [41].

In the regression models, we have treated the observed responses as independent, but there is some dependence in estimated weekly infectious contact ratios over time. Calculating Newey-West heteroskedasticity and autocorrelation consistent sandwich estimators for the parameter variance matrices in the regression models deflated the confidence intervals for the predicted responses slightly in some cases, and increased them slightly in others, but the difference to confidence intervals for predicted responses assuming independence was small.

In summary, we have predicted infectious contact from season, population mask wearing, and mobility, in the specific context of the island of Ireland. Our study corroborates the body of literature demonstrating the effectiveness of mask-wearing [48, 49]. We found that increasing mask-wearing to 90% throughout early 2020 would have decreased inter-individual infectious contact during the first wave, and, in consequence, led to substantially fewer predicted hospitalisations. This effect is likely to have been at least partly due to changes in other factors that mitigate the spread of the virus and were also changing over time, and may be better interpreted as an effect of many accumulated actions to prevent the spread of the virus that were changing over time. Mask-wearing was the one NPI for which international public health bodies and Western governments changed their recommendations; as we have previously noted, early advice strongly discouraged mask-wearing, even claiming this put mask-wearers at higher risk of becoming infected; however by the summer of 2020 the consensus was that mask wearing was beneficial. Changes in public health recommendations about mask wearing likely played a role in the patterns of mask-wearing uptake, leading to very large increases in mask-wearing between March and August 2020; this is in contrast to data on other NPI where the public health messaging was more consistent and changes in behaviour were not as large. It is unlikely that the smaller changes in other NPI could account for the entire effect of mask-wearing we have reported.

## Data Availability

All data produced in the present study are available upon reasonable request to the authors.

