## Supporting Information for "Understanding the role of mask-wearing during COVID-19 on the island of Ireland"

### S1: Mobility Data

We downloaded Google mobility streams for the ROI and the United Kingdom from the Google COVID-19 Community Mobility Reports at [1]. For the overall google mobility stream, we calculated a seven-day moving average of the three streams, transit stations (eg bus and train stations), retail and recreation (eg restaurants, cafes, shopping centres, theme parks, museums, libraries, cinemas) and workplaces. As there were no mobility streams at the level of UK country, we calculated weighted averages of each mobility stream over the 11 subregions of NI, with weights the estimated populations of the subregions, using the mid-2020 dataset from [2].

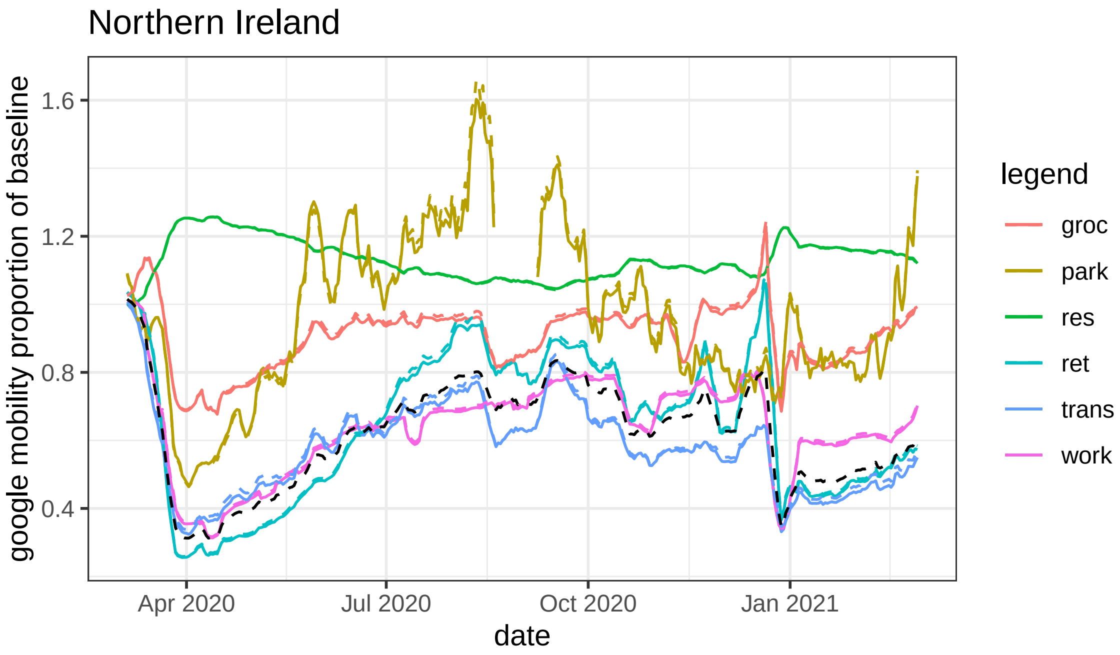


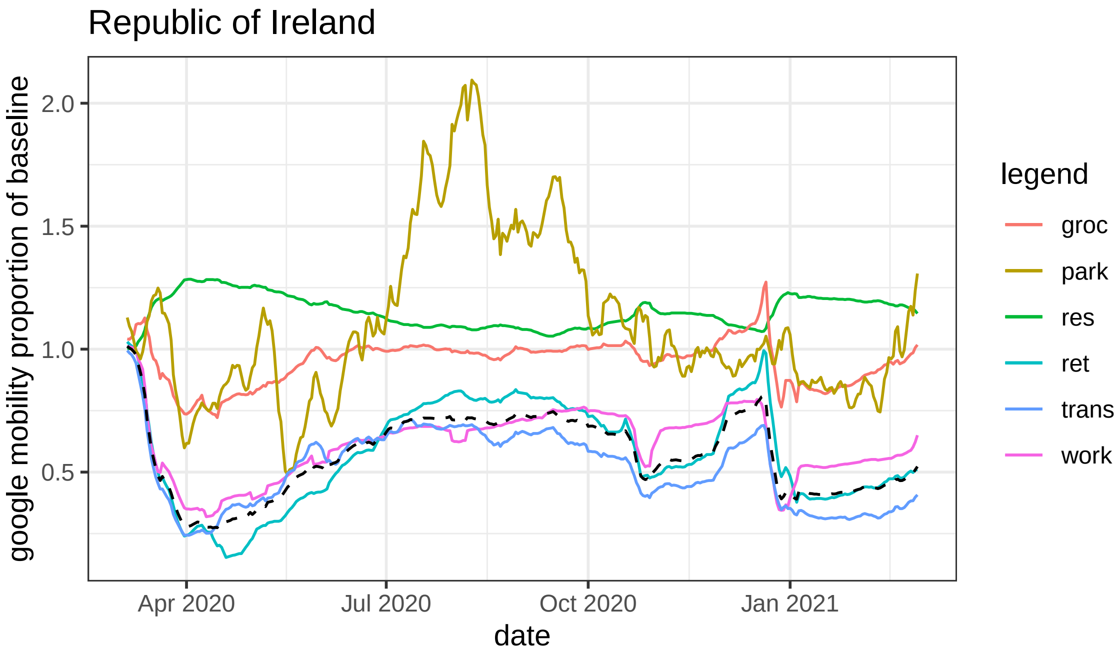


Figure S1: Google mobility streams for the Republic of Ireland and Northern Ireland. We use an average of the workplaces, transit stations and retail and recreation streams as a measure of mobility outside the home - this is shown as a black dashed line in the figure.

### S2: Newey-West confidence bands

[snip]

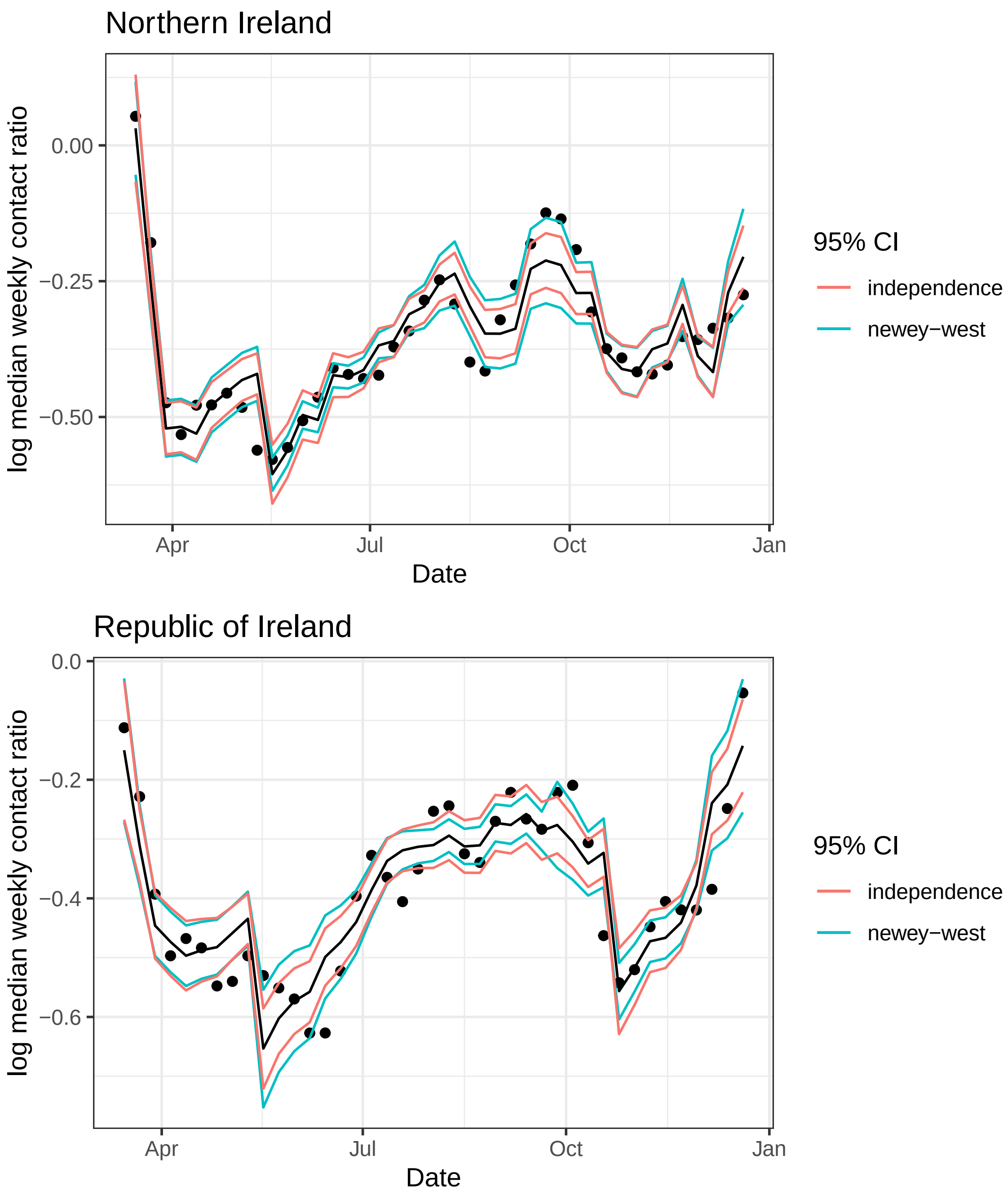


Figure S2: 95% pointwise confidence intervals for predicted responses from linear regression models, comparing the usual independence standard errors with Newey-West standard errors.

### S3: Safe prediction

Extrapolating model predictions beyond observed data means putting a lot of faith in the model, which may be misplaced. Figure S3 shows the regions where there are and are not observed data; predicting the infectious contact ratio in these regions is questionable. For instance, there are no observed data where mobility is close to the baseline value (ie proportion change from baseline is near 1) so attempting to predict from this model the infectious contact ratios with different levels of population mask wearing is not safe. On the other hand, predicting contact ratios when mask wearing is high and mobility changes between about 40% and 80% of the baseline value is safer, being within the range of the observed data.

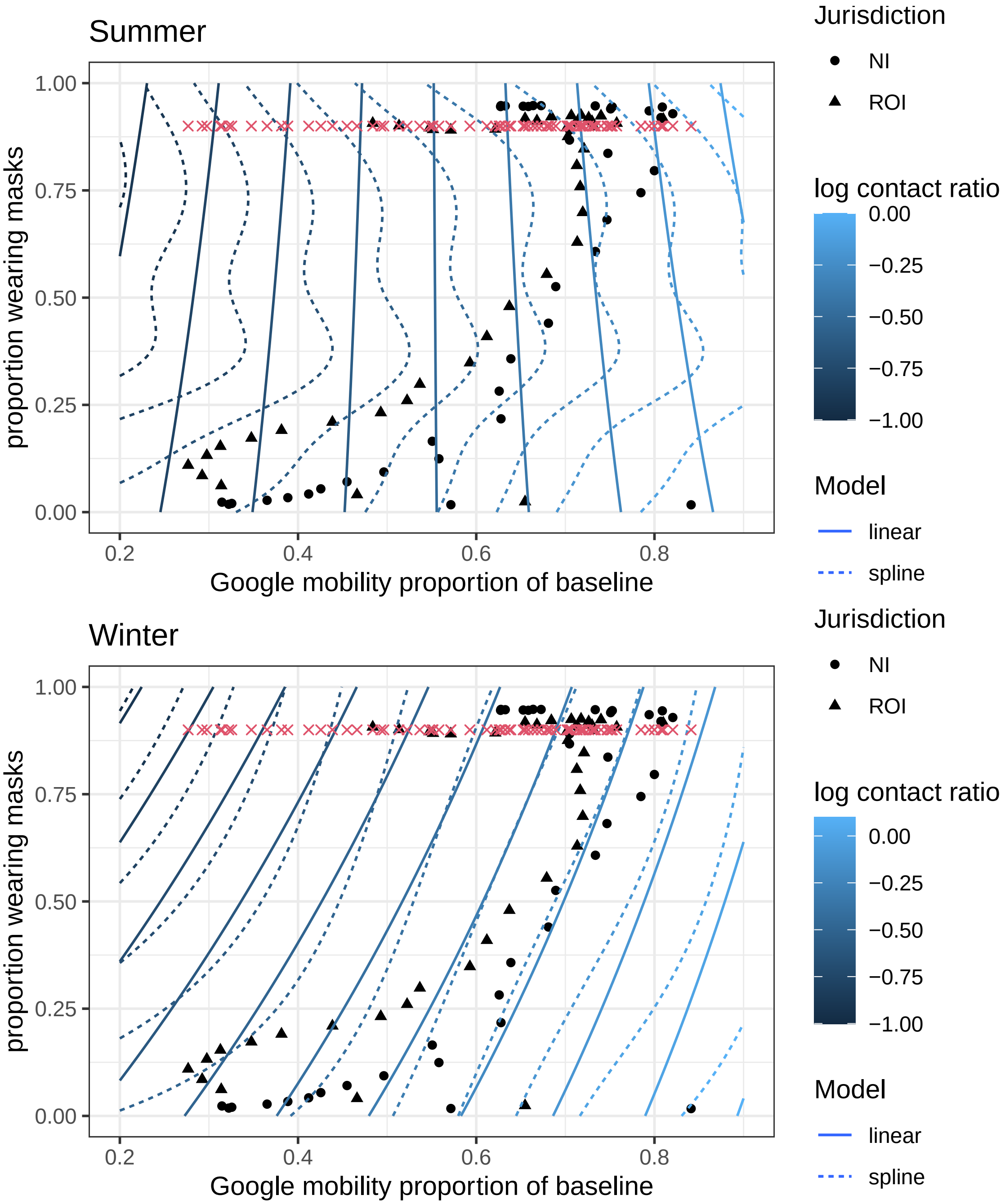


Figure S3: Contour plots for predicted log infectious contact ratio during summer and winter as a function of the mobility and mask variables. The contours show where the predicted log infectious contact ratio is constant. Darker contour lines correspond to lower predicted values. The black points are the observed combinations of mobility and mask variables; extrapolating the model far beyond the observed regions is inadvisable. The red crosses show the hypothetical intervention 90% mask wearing.

[2] Office for National Statistics. Estimates of the population for the UK, England and Wales, Scotland and Northern Ireland;. Available from: https://www.ons.gov.uk/peoplepopulationandcommunity/populationandmigration/populationestimates/datasets/populationestimatesforukenglandandwalesscotlandandnorthernireland.
